# Knowledge and Compliance with Standard Precautions for Nosocomial Infection Prevention Among Undergraduate Nursing and Midwifery Students in Ghana

**DOI:** 10.64898/2026.07.07.26357431

**Authors:** Catherine Tenewaa Osei, Agnes Opoku Asare, Yvonne Oti-Agyen, Hilda Adwubi Osei, Philemon Amooba

## Abstract

**Background:** Healthcare-associated infections (HAIs) represent a significant patient safety challenge in sub-Saharan Africa, where gaps in infection prevention and control (IPC) practice remain prevalent. Nursing and midwifery students are particularly vulnerable during clinical training, yet evidence on their IPC knowledge and compliance in Ghana is limited.

**Objective:** To assess the knowledge of nosocomial infections and the degree of compliance with standard precautions among third-year nursing and midwifery students at Kwame Nkrumah University of Science and Technology (KNUST), Ghana.

**Methods:** A descriptive cross-sectional study was conducted between 28 June and 9 July 2021 at KNUST, Kumasi, Ghana. A total of 150 third-year students (65 nursing, 85 midwifery) completed a structured, self-administered questionnaire adapted from WHO and CDC guidelines. Knowledge was assessed using a 19-item binary scale and compliance using a 17-item Likert-type scale. Respondents were categorised into knowledge and compliance groups using predefined score thresholds for analytical purposes. Chi-square tests, Fisher’s exact test, and Spearman’s rank correlation were used to examine associations between knowledge and compliance.

**Results:** Overall, 143 respondents (95.3%) were categorised as having high knowledge of nosocomial infections and standard precautions (mean score: 16.44/19; SD: 1.59). High compliance with standard precautions was reported by 112 respondents (74.7%; mean score: 59.13/68; SD: 5.89). Compliance was strongest for hand hygiene and glove use but notably lower for PPE use during splash-risk procedures and safe needle-handling practices. No statistically significant association was found between categorised knowledge level and categorised compliance level (χ² = 0.47, df = 1, p = 0.491; Fisher’s exact p = 0.679). When analysed as continuous scores, a modest positive correlation was observed (Spearman’s rho = 0.326, p < 0.001).

**Conclusion:** High knowledge of nosocomial infections did not translate uniformly into high compliance across all standard precaution domains. Practical training, simulation-based learning, and supervised clinical reinforcement are needed to bridge the knowledge–practice gap in nursing and midwifery education in Ghana.

## 1. INTRODUCTION

Healthcare-associated infections (HAIs), also referred to as nosocomial infections, are infections acquired during healthcare delivery that were neither present nor incubating at the time of admission (World Health Organization, 2016). They represent a major global patient safety concern, contributing to increased morbidity, mortality, prolonged hospital stays, and substantial healthcare costs (Allegranzi et al, 2011); (Cassini et al, 2016). The burden of HAIs is disproportionately high in low- and middle-income countries (LMICs), where inadequate infrastructure, limited access to personal protective equipment (PPE), workforce shortages, and systemic gaps in IPC implementation compound the risk (Storr et al, 2017); (Nejad et al, 2011). In sub-Saharan Africa, HAI prevalence estimates have consistently exceeded those reported in high-income settings; some studies report rates approaching three times the global average (Allegranzi et al, 2011; Nejad et al., 2011).

Standard precautions form the cornerstone of evidence-based IPC practice. As defined by the World Health Organization (WHO), standard precautions are a minimum set of infection control practices applied to all patients in all healthcare settings, regardless of diagnosis or presumed infection status (World Health Organization, 2009). They encompass hand hygiene, use of PPE, safe injection practices, respiratory hygiene, safe handling of contaminated equipment, and appropriate waste disposal (World Health Organization, 2009); (Siegel et al, 2007). Consistent adherence to these measures reduces the transmission of healthcare-associated pathogens and protects both patients and healthcare workers (HCWs) (Pittet et al, 2006); (Zingg et al, 2015).

Nursing and midwifery students represent a particularly vulnerable group during clinical training. Their frequent direct patient contact, combined with limited clinical experience, places them at elevated risk of occupational exposure to infectious agents (Prüss-Ustün et al, 2005); (Trim et al, 2003). Students who develop suboptimal IPC habits during training may carry these practices into their professional careers, creating sustained risks for patients and colleagues (Luo et al, 2010). Despite this, the IPC knowledge and compliance behaviours of undergraduate nursing and midwifery students in Ghana have received limited research attention.

Several studies across sub-Saharan Africa have identified persistent gaps between IPC knowledge and actual compliance among HCWs and students (Sahiledengle et al, 2018; Ganczak & Szych, 2007). Factors associated with suboptimal compliance include limited PPE availability, inadequate clinical supervision, high workload, institutional culture, and insufficient reinforcement of IPC protocols during clinical placements (Amoran & Onwube, 2013); (Efstathiou et al, 2011). Understanding the baseline knowledge and compliance behaviours of nursing and midwifery students at a leading Ghanaian university is therefore essential for informing targeted educational interventions and strengthening IPC curricula.

This study aimed to: (i) assess the knowledge of nosocomial infections and standard precautions among third-year nursing and midwifery students at KNUST, Ghana; (ii) describe self-reported compliance with standard precaution practices; and (iii) examine the association between knowledge level and compliance with standard precautions.

## 2. METHODS

### 2.1 Study Design and Setting

A descriptive cross-sectional study was conducted at the Department of Nursing, Faculty of Allied Health Sciences, College of Health Sciences, Kwame Nkrumah University of Science and Technology (KNUST), Kumasi, Ghana. KNUST is a leading public research university in Ghana offering accredited undergraduate programmes in nursing and midwifery. Kumasi is the second-largest city in Ghana and a major hub for healthcare education and service delivery in the Ashanti Region.

### 2.2 Study Population and Sampling

The study population comprised all third-year nursing and midwifery students enrolled at KNUST during the 2020/2021 academic year. Third-year students were selected because they had completed substantial theoretical IPC coursework and had accumulated significant clinical practice experience, making them appropriate participants for assessing both knowledge and self-reported compliance. A census approach was used; all eligible students who provided informed consent were enrolled. A total of 150 students participated (65 nursing students; 85 midwifery students).

### 2.3 Data Collection Instrument

Data were collected using a structured, self-administered questionnaire adapted from validated tools developed by the WHO and the Centers for Disease Control and Prevention (CDC) for assessing IPC knowledge and compliance (World Health Organization, 2009); (Centers for Disease Control and Prevention, 2016). The instrument comprised three sections:

- **Section 1 – Demographic Data:** Age, sex, programme of study, and marital status.

- **Section 2 – Knowledge of Nosocomial Infections and Standard Precautions:** Nineteen binary (Yes/No) items assessing knowledge of nosocomial infection risk factors, the scope and application of standard precautions, hand hygiene indications, glove use indications, and appropriate PPE use during splash-risk procedures.

- **Section 3 – Compliance with Standard Precautions:** Seventeen items assessing self-reported frequency of compliance using a five-point Likert scale (Always, Usually, Sometimes, Seldom, Never), covering hand hygiene, glove use across clinical scenarios, PPE use, and sharps safety practices.

### 2.4 Data Analysis

Data were entered, cleaned, and analysed using IBM SPSS Statistics version 20. Descriptive statistics, including frequencies, percentages, means, and standard deviations, were used to summarise demographic characteristics, knowledge scores, and compliance scores.

**Knowledge scoring:** Knowledge scores were computed from questionnaire items assessing knowledge of nosocomial infections, standard precautions, hand hygiene, glove use, and personal protective equipment. Correct responses were assigned one point and incorrect responses were assigned zero points. Individual scores were summed to obtain an overall knowledge score. Respondents were categorised into knowledge groups using predefined score thresholds for analytical purposes.

**Compliance scoring:** Compliance with standard precautions was assessed using self-reported practice items. Responses were scored according to frequency of compliance. Negatively framed items—specifically the item assessing needle recapping after use—were reverse coded so that higher scores consistently represented safer practice. Individual item scores were summed to obtain an overall compliance score. Respondents were categorised into compliance groups using predefined score thresholds for analytical purposes.

**Missing data:** Missing responses were handled using available-case analysis, whereby respondents were included in analyses for variables with available data.

**Association analysis:** Chi-square tests were used to examine the association between categorised knowledge level and categorised compliance level. Where expected cell frequencies were small, Fisher’s exact test was applied. Spearman’s rank correlation coefficient was used to examine the relationship between continuous knowledge and compliance scores. A p-value of <0.05 was considered statistically significant.

### 2.5 Ethical Considerations

Ethical approval for this study was obtained from the Committee on Human Research and Publication Ethics (CHRPE), School of Medicine and Dentistry, Kwame Nkrumah University of Science and Technology, Kumasi, Ghana.

## 3. RESULTS

### 3.1 Demographic Characteristics

A total of 150 third-year nursing and midwifery students participated in the study. The majority were midwifery students (n = 85; 56.7%) and female (n = 127; 84.7%). Most participants were aged 19–24 years (n = 131; 87.3%) and single (n = 137; 91.3%). Demographic characteristics are summarised in Table 1.

**Table 1.**
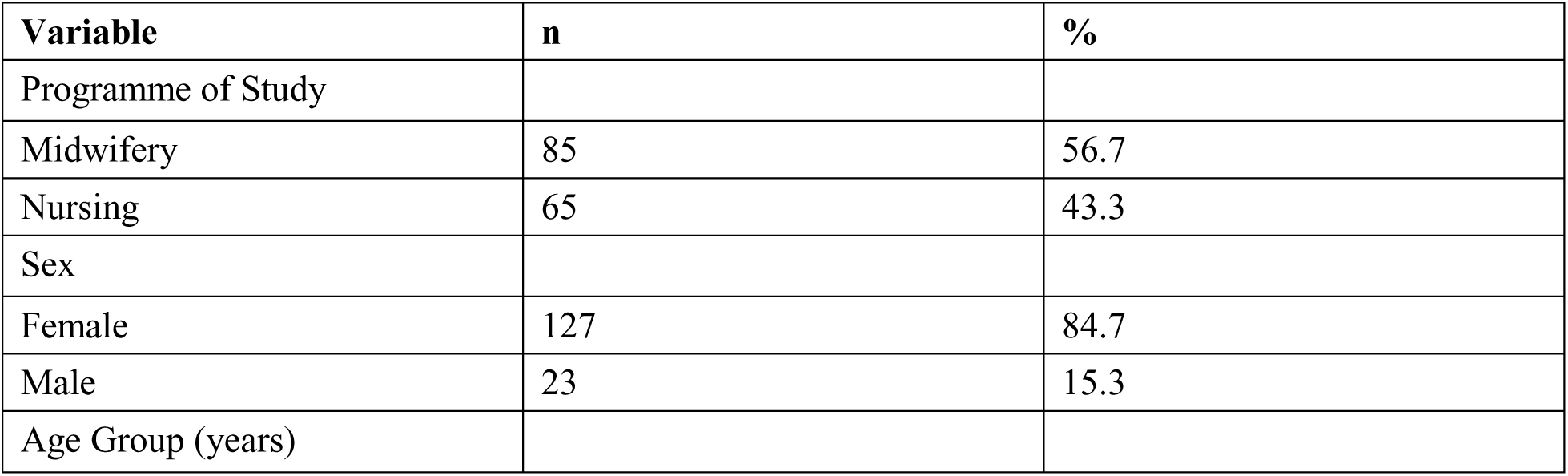
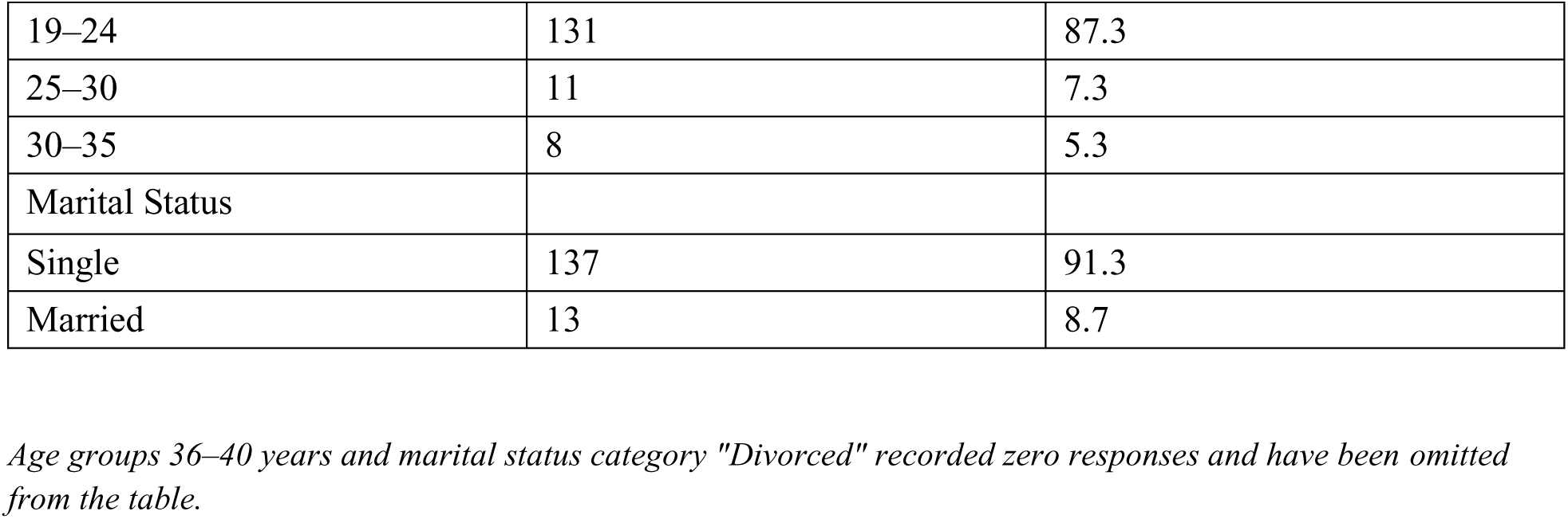
Demographic Characteristics of Participants (N = 150)

### 3.2 Knowledge of Nosocomial Infections and Standard Precautions

The mean knowledge score was 16.44 out of a maximum of 19 points (SD = 1.59; median = 17; range: 11–19). Overall, 143 respondents (95.3%) were categorised as having high knowledge, while seven (4.7%) were classified as having poor knowledge.

Knowledge of nosocomial infection risk factors was strong. Nearly all respondents correctly identified that the environment is a major source of bacteria responsible for nosocomial infections (n = 145; 96.7%), that invasive procedures increase infection risk (n = 144; 96.0%), and that advanced or very young age heightens susceptibility (n = 141; 94.0%).

Regarding the scope of standard precautions, 99.3% (n = 149) correctly recognised that these measures protect both patients and healthcare workers, and 96.0% (n = 144) understood that standard precautions apply to all patients. However, notable misconceptions were identified: 61.3% (n = 92) incorrectly endorsed the view that standard precautions apply only to healthcare workers in contact with body fluids, and 30.0% (n = 45) incorrectly believed that standard precautions protect only patients.

Knowledge of hand hygiene indications was high, with 98.0% (n = 147) correctly identifying the need for hand hygiene after glove removal and 96.0% (n = 144) between patient contacts. Knowledge of glove use indications was similarly strong; 98.0% (n = 147) correctly identified the need for gloves when a healthcare worker has a cutaneous lesion and 96.0% (n = 144) when there is risk of a cut. However, only 69.3% (n = 104) correctly identified the need to wear gloves for each procedure. For PPE use during procedures with splash or spray risk, 94.0% (n = 141) correctly identified the requirement to wear mask, goggles, and gown together. Detailed knowledge responses are presented in Table 2.

**Table 2.** Knowledge of Nosocomial Infections and Standard Precautions (N = 150)

| **Knowledge Item** | **Correct Response** | **Yes n (%)** | **No n (%)** |
| --- | --- | --- | --- |
| Nosocomial Infection Risk Factors |  |  |  |
| Environment (air, water, inert surfaces) is a major source of bacteria | Yes | 145 (96.7) | 5 (3.3) |
| Advanced or very young age increases risk | Yes | 141 (94.0) | 9 (6.0) |
| Invasive procedures increase risk | Yes | 144 (96.0) | 6 (4.0) |
| Scope of Standard Precautions |  |  |  |
| Protects only patients <sup>a</sup> | No | 45 (30.0) | 105 (70.0) |
| Protects patients AND healthcare workers | Yes | 149 (99.3) | 1 (0.7) |
| Applies to all patients | Yes | 144 (96.0) | 6 (4.0) |
| Applies only to HCWs in contact with body fluid <sup>a</sup> | No | 92 (61.3) | 58 (38.7) |
| Hand Hygiene Indications |  |  |  |
| Before or after patient contact | Yes | 133 (88.7) | 17 (11.3) |
| Between patient contacts | Yes | 144 (96.0) | 6 (4.0) |
| After removal of gloves | Yes | 147 (98.0) | 3 (2.0) |
| Glove Use Indications |  |  |  |
| For each procedure | Yes | 104 (69.3) | 46 (30.7) |
| When risk of contact with blood or body fluid | Yes | 138 (92.0) | 12 (8.0) |
| When risk of a cut | Yes | 144 (96.0) | 6 (4.0) |
| When healthcare worker has a cutaneous lesion | Yes | 147 (98.0) | 3 (2.0) |
| PPE Use When Risk of Splashes or Spray |  |  |  |
| Only mask <sup>a</sup> | No | 26 (17.3) | 124 (82.7) |
| Only eye protection <sup>a</sup> | No | 12 (8.0) | 138 (92.0) |
| Only gown <sup>a</sup> | No | 30 (20.0) | 120 (80.0) |
| Mask, goggles, AND gown (full PPE) | Yes | 141 (94.0) | 9 (6.0) |

HCW = healthcare worker; PPE = personal protective equipment.
<sup>a</sup> For items marked with <sup>a</sup>, "No" represents the correct response. "Yes" responses indicate a misconception.

**Table 3.**
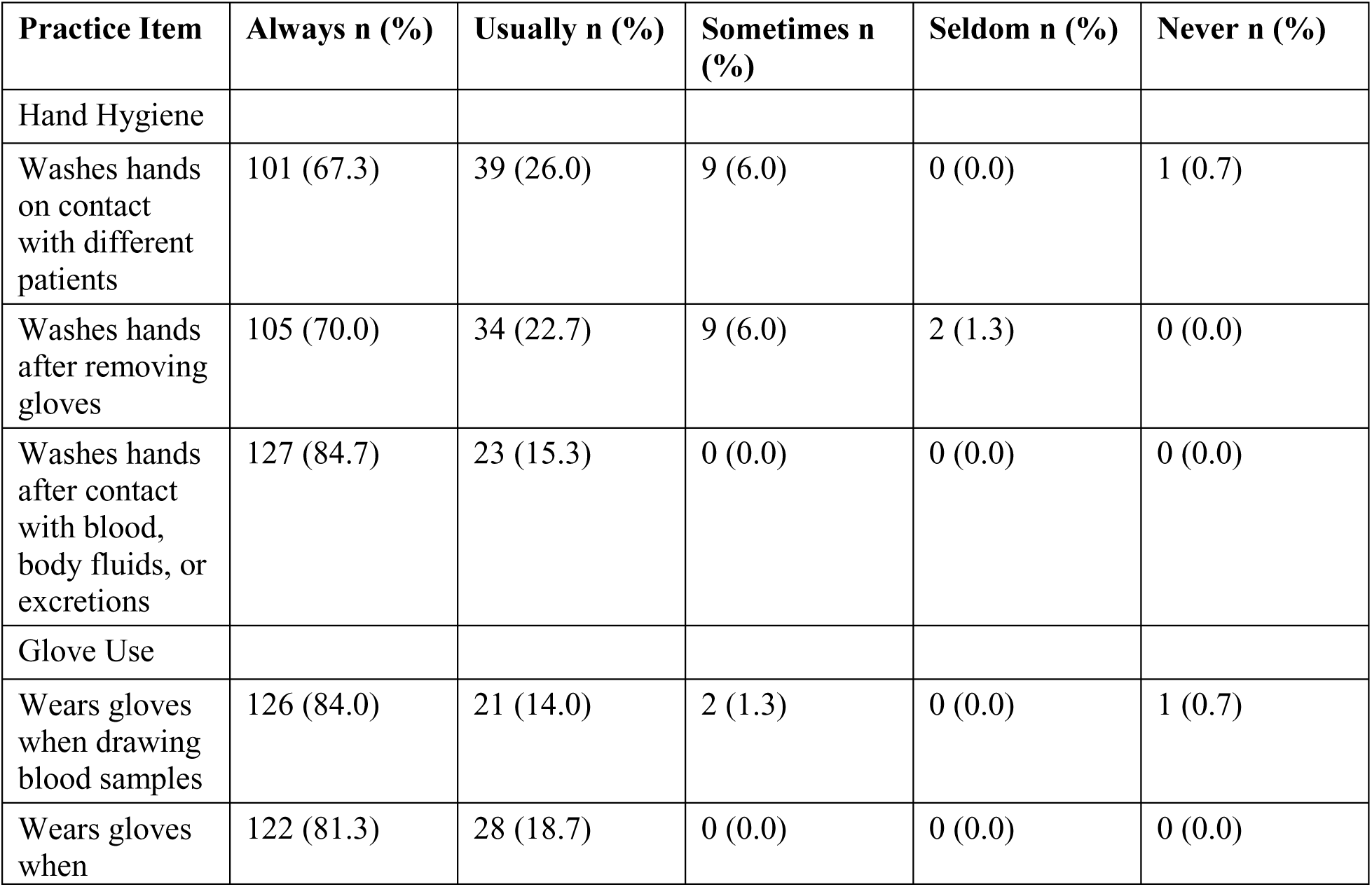
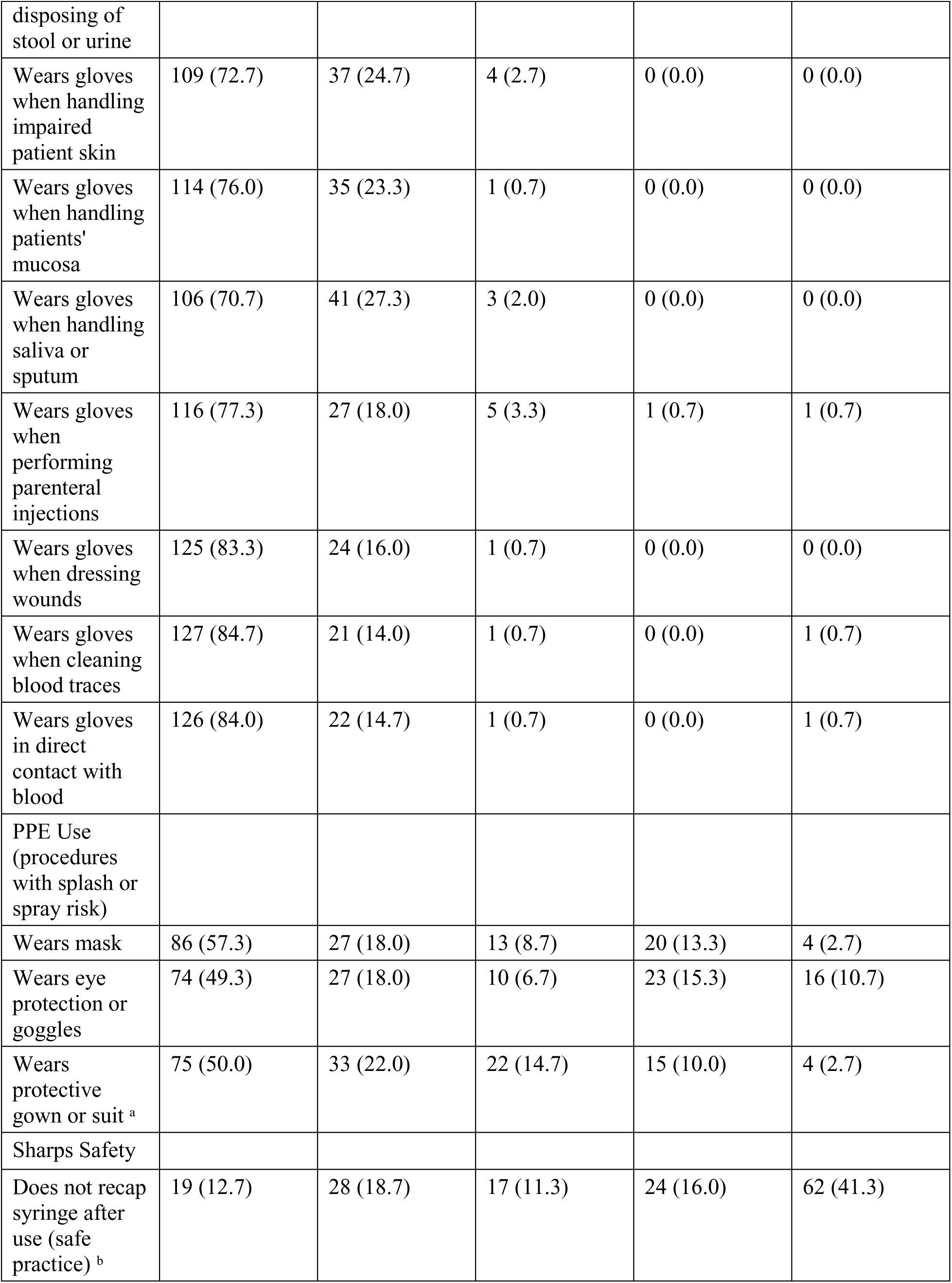
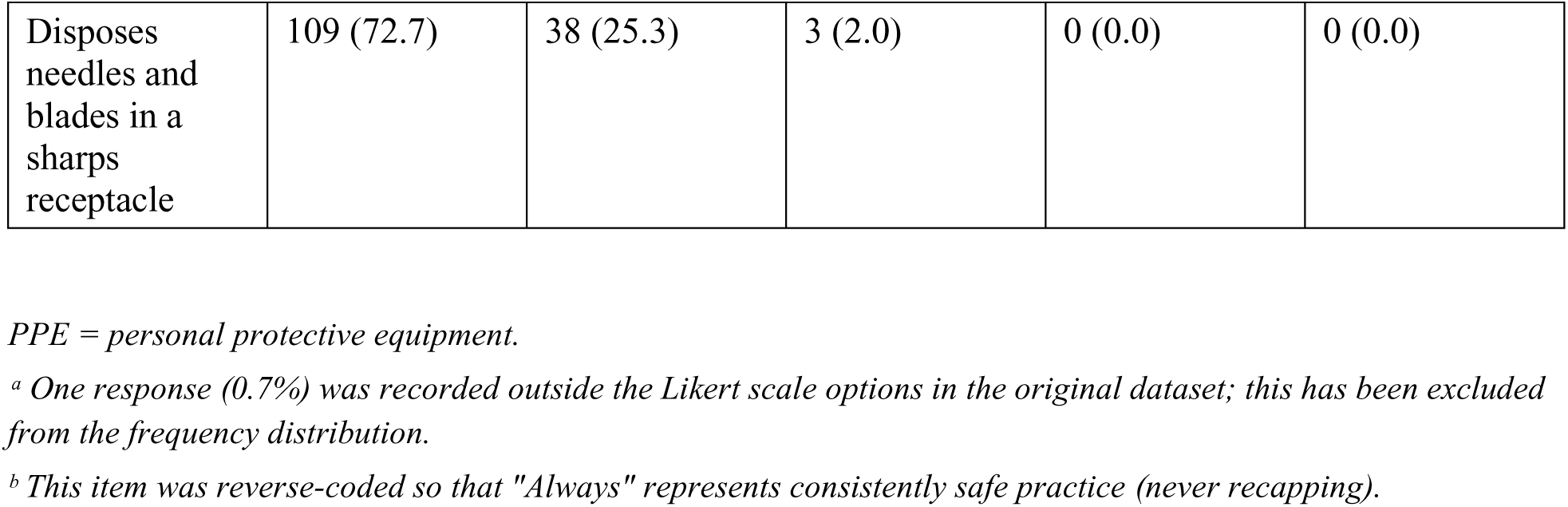
Self-Reported Compliance with Standard Precautions (N = 150)

### 3.3 Compliance with Standard Precautions

The mean compliance score was 59.13 out of a maximum of 68 points (SD = 5.89; median = 60; range: 47–68). High compliance was reported by 112 respondents (74.7%), while 38 respondents (25.3%) were classified as having low compliance.

**Hand hygiene.** Compliance with hand hygiene was generally high. The majority of respondents reported always washing their hands immediately after contact with blood, body fluids, secretions, or excretions (n = 127; 84.7%), and 70.0% (n = 105) always washed hands after removing gloves. Hand hygiene on contact with different patients was always practised by 67.3% (n = 101), with a further 26.0% (n = 39) reporting they usually did so.

**Glove use.** Compliance with glove use was strong across most clinical indications. The highest rates of consistent glove use were observed when cleaning blood traces (n = 127; 84.7%), drawing blood samples (n = 126; 84.0%), and making direct contact with blood (n = 126; 84.0%). Glove use when dressing wounds was always practised by 83.3% (n = 125), and when disposing of stool or urine by 81.3% (n = 122).

**PPE use during splash-risk procedures.** Compliance with full PPE use during procedures with risk of splashes or spray was substantially lower than for hand hygiene and glove use. Only 57.3% (n = 86) reported always wearing a mask, 50.0% (n = 75) always wearing a protective gown, and 49.3% (n = 74) always wearing eye protection during such procedures. Notably, 10.7% (n = 16) reported never wearing eye protection and 13.3% (n = 20) seldom wore masks during these procedures.

Sharps safety. Compliance with safe sharps practices was mixed. Sharps disposal adherence was high, with 72.7% (n = 109) always disposing of needles and blades in a sharps receptacle and 25.3% (n = 38) usually doing so. However, consistent safe needle non-recapping behaviour was less common: after reverse coding, only 12.7% (n = 19) always avoided recapping syringes after use.

### 3.4 Association Between Knowledge and Compliance

Chi-square analysis was performed to assess the association between categorised knowledge level and categorised compliance level. Although the majority of respondents demonstrated both high knowledge and high compliance, no statistically significant association was found between the two categorical variables (χ² = 0.47, df = 1, p = 0.491). Fisher’s exact test yielded a consistent result (p = 0.679), confirming the absence of a statistically significant association.

When analysed as continuous scores, a modest positive correlation was observed between knowledge and compliance (Spearman’s rho = 0.326, p < 0.001), indicating that respondents with higher knowledge scores tended to report higher compliance with standard precautions.

|  | High Compliance n (%) | Low Compliance n (%) | Total n |
| --- | --- | --- | --- |
| High Knowledge (n = 143) | 107 (74.8) | 36 (25.2) | 143 |
| Poor Knowledge (n = 7) | 5 (71.4) | 2 (28.6) | 7 |
| Total | 112 (74.7) | 38 (25.3) | 150 |

$\chi^2 = 0.47$ , $df = 1$ , $p = 0.491$ ; Fisher's exact test $p = 0.679$ .
Spearman's $\rho = 0.326$ , $p < 0.001$ (continuous knowledge and compliance scores).

## 4. DISCUSSION

This study assessed the knowledge of nosocomial infections and self-reported compliance with standard precautions among third-year nursing and midwifery students at KNUST, Ghana. The principal findings were: (i) the majority of respondents demonstrated high knowledge of nosocomial infections and standard precautions; (ii) overall compliance was high in most domains, but notable gaps were identified in PPE use during splash-risk procedures and safe needle-handling practices; and (iii) no statistically significant association was found between categorised knowledge level and categorised compliance, although a modest positive correlation was observed between continuous knowledge and compliance scores.

### 4.1 Knowledge of Nosocomial Infections and Standard Precautions

The high level of knowledge observed in this study (95.3% categorised as high knowledge; mean score: 16.44/19) is consistent with findings from comparable studies among some African settings, including Nigeria (Abdulraheem et al, 2012). However, studies from other settings have reported lower levels of infection-prevention knowledge and practice, highlighting considerable variability across healthcare environments (Sahiledengle et al, 2018). This may reflect the integration of IPC content within the nursing and midwifery curriculum at KNUST, as well as the heightened awareness of infection prevention practices that characterised the clinical environment during the period in which this study was conducted (World Health Organization, 2021).

Despite overall high knowledge scores, specific misconceptions were identified. Notably, 61.3% of respondents incorrectly believed that standard precautions apply only to healthcare workers in contact with body fluids, and 30.0% incorrectly believed they protect only patients. These misconceptions are clinically significant: a misunderstanding of the universal applicability of standard precautions may lead to selective and inconsistent implementation in clinical practice. Similar conceptual gaps have been reported in other African settings and may reflect areas where curriculum emphasis or clinical reinforcement is insufficient. Additionally, only 69.3% correctly identified the need to wear gloves for each procedure, suggesting that some students may apply a risk-based rather than universal approach to glove use.

### 4.2 Compliance with Standard Precautions

Although overall compliance was high (74.7% classified as high compliance; mean score: 59.13/68), the pattern of compliance was uneven across practice domains. Hand hygiene and glove use were performed consistently by the majority of respondents, a pattern that is broadly comparable with reports from other African healthcare settings where adherence to core standard precautions varies by practice domain (Amoran & Onwube, 2013; Haile et al., 2017; Desta et al., 2018). The high rate of hand hygiene after contact with blood and body fluids (84.7% always) and consistent glove use when drawing blood or cleaning blood traces (84.0–84.7% always) suggests that students have internalised these core practices, likely reinforced through repeated clinical exposure and direct supervision.

In contrast, compliance with PPE use during procedures with splash or spray risk was substantially lower. Only approximately half of respondents reported always wearing eye protection (49.3%) or a protective gown (50.0%), and 10.7% reported never wearing eye protection at all. These findings are consistent with those of Amoran and Onwube (Amoran & Onwube, 2013), Desta et al. (Desta et al, 2018), and other investigators who have documented persistently low PPE compliance among healthcare workers and students in resource-limited settings, often attributed to limited PPE availability, discomfort, time pressures, and insufficient clinical supervision (Amoran & Onwube, 2013); (Tenna et al, 2013). It is also plausible that access to certain PPE items—particularly eye protection and gowns—may have been constrained in the training environment during the study period, a contextual factor that should be acknowledged when interpreting these findings.

Safe needle-handling practices also revealed a gap. After reverse coding, only 12.7% of respondents consistently avoided recapping syringes, a practice associated with a substantial proportion of needlestick injuries among healthcare workers globally (Prüss-Ustün et al, 2005); (Mbaisi et al, 2013). This finding is concerning given that needlestick injuries represent a major occupational hazard for nursing and midwifery students during clinical training. In contrast, sharps disposal compliance was high (72.7% always), suggesting that students have retained training on appropriate waste disposal.

### 4.3 The Relationship Between Knowledge and Compliance

The absence of a statistically significant association between categorised knowledge level and categorised compliance (χ² = 0.47, p = 0.491) suggests that possessing high theoretical knowledge of standard precautions does not, in itself, guarantee consistent adherence in clinical practice. This finding is consistent with those of multiple studies examining the knowledge–practice gap in IPC among healthcare workers and students in Africa and beyond (Sahiledengle et al, 2018); (Ider et al, 2012); (Sax et al, 2007).

Although categorised knowledge level was not significantly associated with categorised compliance level, continuous score analysis demonstrated a modest positive correlation between knowledge and compliance (Spearman’s rho = 0.326, p < 0.001). This finding suggests that increased knowledge may contribute to better infection prevention practices, although additional factors beyond knowledge are likely to influence adherence to standard precautions. The discrepancy between the categorical and continuous analyses underscores the importance of how variables are operationalised in IPC research and suggests that future studies should consider reporting both categorical and continuous measures.

The high level of knowledge observed among participants may reflect the infection prevention and control content embedded within the nursing and midwifery curriculum, as well as repeated exposure to clinical training environments. Nevertheless, the persistence of suboptimal compliance in selected domains suggests that knowledge alone is insufficient to ensure consistent adherence to standard precautions. Behavioural, environmental, and institutional factors—including PPE availability, clinical supervision, workload, and institutional culture—may influence practice independently of knowledge levels (Amoran & Onwube, 2013; Efstathiou et al, 2011). Educational interventions should therefore combine theoretical instruction with simulation-based training, structured supervision, and continuous reinforcement of infection prevention practices during clinical placements.

### 4.4 Implications for Nursing and Midwifery Education

The findings of this study have important implications for nursing and midwifery education in Ghana and similar low-resource settings. The identification of specific knowledge gaps, particularly regarding the universal application of standard precautions and the appropriate use of full PPE points to areas where curriculum content and pedagogical approaches may benefit from targeted revision.

The findings highlight the importance of integrating infection prevention and control training throughout nursing and midwifery curricula, rather than confining it to discrete theoretical modules. Regular practical training, competency assessments, simulation exercises, and supervised clinical experiences may help translate theoretical knowledge into consistent adherence to standard precautions during clinical placements (Cant & Cooper, 2010); (Bello et al, 2011). Simulation-based learning approaches, which allow students to practise IPC procedures in safe and controlled environments, have demonstrated effectiveness in improving both knowledge and clinical performance in IPC and should be considered as a complement to didactic instruction (Cant & Cooper, 2010).

At the institutional level, healthcare facilities hosting nursing and midwifery students should ensure adequate availability of PPE and sharps safety equipment, provide structured IPC orientation for incoming students, and establish systems for regular compliance monitoring and feedback. Clinical supervisors and preceptors play a critical role in modelling and reinforcing correct IPC practices and should themselves receive ongoing IPC training and support (Efstathiou et al, 2011); (Haile et al, 2017).

At the national level, the findings support advocacy for the inclusion of standardised IPC competency assessments within the licensing and registration requirements for nurses and midwives in Ghana, consistent with WHO global IPC minimum requirements (World Health Organization, 2009); (Ministry of Health, Ghana, 2009).

### 4.5 Limitations

Several limitations of this study should be considered when interpreting the findings. First, this study relied exclusively on self-reported compliance measures, which may be subject to recall bias and social desirability bias. Consequently, actual compliance practices may differ from reported behaviours; observational methods would provide a more objective assessment and should be considered in future research. Second, the cross-sectional design limits causal inferences regarding the relationship between knowledge and compliance; it is not possible to determine from these data whether improvements in knowledge lead to improvements in compliance. Third, the study was conducted at a single institution, which limits the generalisability of findings to other nursing and midwifery training institutions in Ghana or elsewhere in sub-Saharan Africa. Fourth, the questionnaire was not formally validated in the Ghanaian context, which may affect the reliability of the knowledge and compliance scales.

Fifth, because the original scoring criteria used during the undergraduate thesis analysis were unavailable, knowledge and compliance scores were reconstructed from the original dataset using transparent and reproducible scoring procedures. Although this approach enabled secondary analysis of the data, slight differences from the original scoring framework cannot be excluded.

Finally, the data were collected in 2021 during the COVID-19 pandemic, a period characterized by heightened infection prevention and control awareness and reinforcement. Consequently, compliance levels observed in this study may not fully reflect current post-pandemic practices among nursing and midwifery students.

## 5. CONCLUSION

This study found that the majority of third-year nursing and midwifery students at KNUST, Ghana, demonstrated high knowledge of nosocomial infections and standard precautions and reported high overall compliance with standard precaution practices. However, notable compliance gaps were identified, particularly in the use of full PPE during procedures with splash or spray risk and in safe needle-handling practices. No statistically significant association was found between categorised knowledge level and categorised compliance, although a modest positive correlation was observed between continuous knowledge and compliance scores. These findings underscore the importance of moving beyond theoretical instruction in IPC education to emphasise practical skill development, simulation-based training, and supervised clinical reinforcement. Strengthening IPC training within nursing and midwifery curricula, ensuring adequate PPE availability during clinical placements, and implementing competency-based assessments are recommended to reduce the burden of healthcare-associated infections in Ghana and similar settings.

## DECLARATIONS

### Ethics approval and consent to participate

Ethical approval was granted by the Committee on Human Research and Publication Ethics (CHRPE), School of Medicine and Dentistry, Kwame Nkrumah University of Science and Technology, Kumasi, Ghana. Written informed consent was obtained from all participants.

## Consent for publication

Not applicable.

## Authors’ Contributions

Catherine Tenewaa Osei conceptualized the study, conducted the literature review, participated in instrument development and data collection, performed data analysis and interpretation, prepared the original manuscript draft, and led manuscript revision and preparation for publication. Agnes Opoku Asare contributed to data collection, review of study materials, and critical review of the manuscript. Yvonne Oti-Agyen contributed to data collection, review of study materials, and critical review of the manuscript. Hilda Adwubi Osei participated in literature review, contributed to data analysis and critical review of the manuscript. Philemon Amooba provided academic supervision, methodological guidance, and critical review of the manuscript. All authors reviewed and approved the final manuscript prior to submission.

## Data Availability Statement

The dataset supporting the findings of this study is available from the corresponding author upon reasonable request.

## Data Availability

All data produced in the present study are available upon reasonable request to the authors.

